# A spatio-temporal mapping and bayesian modelling of risk factors of pneumonia symptoms in under-five children in Nigeria

**DOI:** 10.1101/2022.12.19.22283675

**Authors:** K. A. Atoloye, T. V. Lawal, A. S. Adebowale, A.F. Fagbamigbe

## Abstract

**Background:** Pneumonia remains a public health challenge in most parts of the world, with Nigeria having the highest number of pneumonia-related deaths. Understanding the geographical distribution, trends and risk factors associated with pneumonia symptoms will aid an appropriate intervention of pneumonia and subsequently reduce its burden in Nigeria.

**Method:** This cross-sectional study used data from the 2008, 2013 and 2018 Nigeria Demographic Health Survey. In each of the survey round, a multi-stage cluster sampling technique was used to select the eligible respondents who are women of reproductive age. The outcome variables are the presence of key symptoms of pneumonia: fever, cough, and short rapid breaths. Optimized hotspot analysis was used to identify states with a significantly high prevalence of pneumonia symptoms, MCMC mixed-effect models were fitted to each symptom.

**Results:** The prevalence of cough was 12.1%, 10.1% and 16.9% in 2008, 2013 and 2018 respectively, 16.2%, 13.3%, and 25.7% for fever; and 41.7%, 42.5% and 6.5% for short rapid breaths respectively with variations across the states. The adjusted odds of having a cough among the children aged 6-11, 12-23, 24-35 and 36-47 months were 95% higher (adjusted odds ratio (aOR) =1.95, 95% Credible Interval (CrI): 1.77, 2.18), 92% higher (aOR=1.92, 95% CrI: 1.73, 2.12), 45% higher (aOR=1.45, 95% CrI: 1.31, 1.62) and 15% higher (aOR=1.15, 95% CrI: 1.03, 1.27) respectively, relative to ages 0-5 months. Similar patterns were noticed for fever and short rapid breaths. Mothers’ education was significant for cough and fever but not for short rapid breaths. Mothers’ age was significant only for short rapid breaths at higher odds. Other significantly associated factors with symptoms include residence type, housing quality, wealth index and region.

**Conclusion:** Fever, cough, and short rapid Breaths are prevalent among under-five children in Nigeria. These symptoms are associated with different characteristics and varied across states in Nigeria. Therefore, it is pertinent that mothers improve on the available preventive and management strategies with the view to mitigating the consequences of pneumonia symptoms among under-five children in Nigeria.

## Introduction

Pneumonia is an acute respiratory infection in one or both lungs, characterized by cough, fever, shortness of breath, shallow breathing, low energy, etc. [1]. On an annual basis, the disease has accounted for over ten million hospital admissions and more than a million deaths among under-five children globally [2,3]. In 2017, pneumonia was identified as the 4^th^ leading cause of mortality and may eventually become the 3^rd^ by 2024 [4]. According to the World Health Organization (WHO), pneumonia accounts for about 14% of the disease burden globally [2].

In Africa, pneumonia is a potentially life-threatening ailment [2,5,6]. It is not just a major causal factor of mortality and morbidity, but it is as well associated with a non-negligible burden on healthcare infrastructures [7,8] and incomes of households [9]. Oftentimes, pneumonia involves several pathogens that are transmissible among individuals. Analyses have identified various pneumonia results to be temporally seasonal [10,11]. Paynter et al. found that admissions based on pneumonia are clustered spatially on a high level aided by contact with infected individuals [12].

WHO estimated 868,000 deaths as a result of pneumonia in 2010 with 140,000 deaths on an annual basis among under-five children, the highest in Africa [13]. In Nigeria, pneumonia is one of the leading communicable diseases. Nigeria is one of the countries with the highest burden of pneumonia in childhood and accounts for 162,000 pneumonia-related deaths worldwide in 2018, constituting 20.2% of global deaths [14]. The prevalence of pneumonia in a study conducted in Nigeria in 2013 was 13.3% and 23.9% in another study in 2015 [15,16]. This high prevalence may attract poor health outcomes within the population if unchecked. Therefore, it is of utmost importance to abate the prevalence of pneumonia symptoms in Nigeria to ensure not just healthy living in children but for the future national sustainability as emphasized in the Sustainable Development Goal (SDG)-3, to “Ensure healthy lives and promote well-being for all and sundry and to bring to an end, avoidable deaths of under-five children with all countries aiming to abate neonatal deaths to as low as 12 per 1 000 live births and under-5 deaths to as low as 25 per 1 000 live births’ by 2030” [17].

Several factors have been identified as risk factors for childhood pneumonia, ranging from indoor air pollution due to biomass, outdoor air pollution as a result of vehicular emission and many other factors [18,19]. Previous studies have also documented that the air intake from polluting fuel is an influential risk factor for acute respiratory infection among under-five children in developing countries [20–27].

Other studies have shown that some socioeconomic characteristics like maternal education, region of residence, and household wealth status are important precursors to the risk factors for pneumonia symptoms among children [28–33]. Moreover, studies have also demonstrated that rural-urban residence [34], maternal age at the child’s birth and the child’s birth order (Mishra, 2003) and water and sanitation facilities [35] are other risk factors for child pneumonia. However, most of these studies focused on the actual disease but not the individual possible symptoms. Addressing issues around the symptoms is more beneficial to the population’s health than the disease itself.

It is also important to note that most of these earlier studies used only descriptive and traditional frequentist approaches. Concurrent assessment of the geographical distribution, trends and risk factors associated with pneumonia symptoms remains a grey area for research in Nigeria. Understanding the spatial-temporal distribution, trends and risk factors associated with pneumonia symptoms will aid the prevention of pneumonia and subsequently reduce its prevalence and attendant burden. Therefore, this study employed a Markov Chain Monte Carlo Random Intercept Logistic Regression to identify and explain the risk factors associated with pneumonia symptoms (cough, fever and short rapid breaths). This study makes an original contribution to existing knowledge on pneumonia by providing information on spatial-temporal distribution and drivers of the symptoms of pneumonia among under-five children in Nigeria. Our study outcomes will aid public health efforts toward reducing the incidence of pneumonia in children.

## Methods

### Study Area

This study was conducted in Nigeria - a West African country with an estimated population of about 215 million. Nigeria consists of six (6) geo-political zones (North-Central, North-East, North-West, South-East, South-South and South-West), which are further divided into 36 states and the Federal Capital Territory (FCT), where each state consists of a third-tier local government and wards. Temperature and humidity are fairly constant all-round year in the south, while the seasons vary considerably in the north. Nigeria’s climate is relatively hot and is usually classified into two seasons, dry and wet. The dry season runs from November to March, whereas, the wet season begins in April and ends in October. These seasons come with the symptoms of pneumonia depending on individuals’ susceptibility, adaptation, and body reactions to changes in environmental conditions.

### Data Collection

The data used in this study were nationally representative and based on the Nigeria Demographic Health Survey (NDHS) conducted in 2008, 2013 and 2018. In the surveys, the unit of inquiry was women of reproductive age (15 to 49 years). They were asked questions regarding their reproductive health, births and children alongside other related topics. The sampling of the respondents during the survey was based on a 2-stage stratified cluster design (National Population Commission [Nigeria] and ICF International, 2014). Data were collected from all eligible women in selected households with the aid of a pretested questionnaire by trained interviewers. Data collected from women about their under-five children were specially processed to constitute the children’s recode file which was utilized for analysis in this study. A detailed explanation of the field processes has been reported elsewhere [39– 41]

### Derivation of Variables

The outcome variables for this study are fever, cough, and short and rapid breaths as symptoms of pneumonia. Each outcome variable measures the presence of the symptoms as “yes” or “no” with value labels “1” or “0” respectively.

Based on existing literature, the explanatory variables were grouped into household, maternal and children characteristics. The maternal background characteristics are age, education, household wealth quintile, place of residence, and geopolitical zone. Children’s characteristics include current age in months and sex. The household characteristics included cooking fuel (clean/unclean), drinking water source (improved or unimproved), housing material (improved or unimproved) and toilet facility (improved or unimproved). Clean fuel includes electricity, and liquefied natural gas/biogas and unclean fuel include wood, charcoal, kerosene, straw shrubs, animal dung and grass. The improved sources of drinking water include a protected well, borehole, bottled water and spring rainwater. The unimproved includes a spring tanker, unprotected well with drum, sachet water, surface water, and other sources. The housing material quality derivation was based on a composite score of the type of wall, floor and roof materials. If cement/carpet/rug/ceramic tiles/vinyl asphalt strips were used for the floor, the floor quality is coded 1 else it is coded 0. In the same vein, wall material quality is coded 1 is made of cement blocks/bricks else 0. If the roof material is made of calamine/cement roofing shingles/cement fibres/ceramic tiles/zinc, it is coded 1 else 0. Categories of the housing material were further aggregated and coded as poor (1), average (2) and good (3). For toilet facilities, it was coded as improved or unimproved depending on the type of toilet facility used.

### Data Analyses

Descriptive statistics, Bayesian MCMC logistic regression and spatial analyses were used. The spatial variation of the prevalence of the symptoms of pneumonia was analysed using the Optimized Hotspot Analysis. It entails the creation of maps of statistically significant hot and cold spots using the Getis-Ord Gi* statistic employing incident points or weighted features (points or polygons). It evaluates the characteristics of the input feature class to produce optimal results. Optimized Hotspot analysis uses the Getis-Ord Gi* statistic using a fixed distance band in ArcGIS software [42]. The Bayesian MCMC logistic regression model is as specified below:

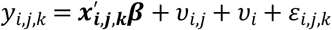

For *k*=1…*n*_*ij*_ level-1 units that are nested within *j*=1…*n*_*i*_ level-2 units that are in turn nested within e *i*=1…*n* level-3 units.

where 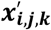 is the covariate vector, ***β*** are unknown regression parameters, *v*_*i*_is the unknown random effect at level 3, *v*_*i,j*_ is the unknown random effect at level-2, and *ε*_*i,j,k*_ are the model residuals that follow a logistic distribution. The model is a random-intercept mixed-effect model. Level 1 is fixed and consists of fixed effect predictors while levels 2 and 3 are the random effect levels defined by the group-level variables. Level 2 group level is defined by the community groups/cluster group variable while the level 3 group is defined by the state group variable. No group-level predictor is involved as interest is more of the amount of variation contributed by the group-level variables. The absence of group-level predictors makes the model a random-intercept mixed model.

In fitting the MCMC model, parameters specified in the *brms* package in R included 4 chains, 4 cores, 4 threads, 50,000 iterations and a thinning rate of 50. A weakly informative prior, derived from the sample was also used.

## Results

### Distribution of Pneumonia Symptoms and the Demographic Characteristics of the Children and their Mothers

#### Distribution of Cough

Table 1 shows the prevalence of cough between 2008 and 2013. The prevalence of cough is highest in 2018 (16.9%) and lowest in 2013 (10.1%). Across the survey years, the prevalence of cough was consistently highest among children aged 6 to 11 months and least among women with no formal education where the prevalence was 9.9%, 7.6%, and 14.7% in 2008, 2013, and 2018 survey year respectively.

**Table 1.** Prevalence of Cough by Selected Demographic Characteristics, per Year.

|  | **2008**<br>**(N=22139)**<br>**n (%)** | **2013**<br>**(N=26076)**<br>**n (%)** | **2018**<br>**(N=11375)**<br>**n (%)** |
| --- | --- | --- | --- |
| **Total** | 2673 (12.1) | 2630 (10.1) | 1921 (16.9) |
| **<u>Characteristics</u>** |  |  |  |
| **Sex of Child** | **2.71** | **0.887** | **0.052** |
| Male | 1319 (11.7) | 1350 (10.3) | 968 (16.8) |
| Female | 1354 (12.4) | 1280 (9.9) | 953 (17.0) |
| **Child Age (In Months)** | **147.532\*** | **268.016\*** | **92.017\*** |
| 0-5 | 257 (9.7) | 205 (7.3) | 164 (13.6) |
| 6-11 | 399 (15.5) | 429 (14.1) | 271 (22.0) |
| 12-23 | 684 (15.3) | 763 (14.1) | 496 (20.8) |
| 24-35 | 553 (13.4) | 513 (10.5) | 360 (16.5) |
| 36-47 | 424 (9.7) | 407 (8.0) | 362 (16.6) |
| 48-59 | 356 (9.0) | 313 (6.5) | 268 (12.3) |
| **Maternal Age** | **11.237** | **19.614\*** | **17.875\*** |
| 15-19 | 158 (13.4) | 140 (11.3) | 69 (15.3) |
| 20-24 | 524 (12.3) | 566 (11.3) | 388 (19.2) |
| 25-29 | 793 (12.5) | 718 (9.8) | 580 (17.9) |
| 30-34 | 587 (12.3) | 589 (10.2) | 414 (15.6) |
| 35-39 | 370 (11.2) | 383 (9.3) | 302 (15.6) |
| 40-44 | 174 (11.0) | 166 (8.5) | 118 (15.2) |
| 45-49 | 67 (9.7) | 68 (9.4) | 50 (17.0) |
| **Maternal Education** | **99.797\*** | **170.411\*** | **29.459\*** |
| No Education | 1063 (9.9) | 935 (7.6) | 646 (14.7) |
| Primary | 702 (13.5) | 622 (11.5) | 375 (19.8) |
| Secondary | 765 (15.0) | 888 (13.0) | 725 (17.9) |
| Higher | 143 (12.6) | 185 (12.2) | 175 (16.8) |
| **Source of Drinking Water** | **0.008** | **0.019** | **1.283** |
| Improved | 1372 (12.1) | 1561 (10.1) | 1153 (16.6) |
| Unimproved | 1301 (12.1) | 1069 (10.1) | 768 (17.4) |
| **Cooking Fuel Type** | **23.816\*** | **1.991** | **2.481** |
| Clean | 466 (14.7) | 451 (10.7) | 361 (15.8) |
| Unclean | 2207 (11.6) | 2179 (10.0) | 1560 (17.2) |
| **Toilet Facility** | **0.406** | **0.441** | **2.21** |
| Improved | 1295 (12.2) | 1268 (10.0) | 1049 (17.4) |
| Unimproved | 1378 (11.9) | 1362 (10.2) | 872 (16.3) |
| **Housing Quality Material** | **30.726\*** | **5.101** | **14.678** |
| Poor | 1084 (10.8) | 1087 (9.9) | 539 (19.2) |
| Average | 374 (12.2) | 392 (9.4) | 326 (16.7) |
| Good | 1215 (13.4) | 1151 (10.5) | 1056 (16.0) |
| **Place of Residence** | **2.361** | **8.192\*** | **0.701** |
| Urban | 783 (12.6) | 921 (10.9) | 733 (16.5) |
| Rural | 1890 (11.9) | 1709 (9.7) | 1188 (17.1) |
| **Wealth Index** | **36.247\*** | **29.890\*** | **13.817** |
| Poorest | 596 (10.9) | 498 (8.4) | 434 (19.2) |
| Poorer | 560 (10.7) | 614 (10.1) | 362 (15.9) |
| Middle | 544 (12.6) | 605 (11.3) | 419 (16.7) |
| Richer | 537 (13.8) | 489 (10.1) | 408 (17.2) |
| Richest | 436 (13.6) | 424 (11.0) | 298 (15.3) |
| **Region** | **567.619\*** | **745.567\*** | **337.491** |
| North Central | 353 (8.4) | 324 (8.2) | 307 (15.3) |
| North East | 817 (15.5) | 914 (16.7) | 544 (26.7) |
| North West | 346 (6.3) | 362 (4.4) | 304 (10.9) |
| South East | 408 (20.5) | 364 (15.5) | 298 (18.6) |
| South South | 477 (19.8) | 449 (14.5) | 310 (24.7) |
| South West | 272 (9.7) | 217 (7.4) | 158 (9.5) |

**\*Chi-square values significant at 95% C.I.**

#### Distribution of Fever

Table 2 also shows that the prevalence of fever dipped between 2008 and 2013, from 16.2% to 13.3% and rose to 25.8% in 2018. Across the survey years, the data show that children in the age group 12-23 months mostly experienced fever more than their counterparts in the other age group while the least prevalence was found among those aged 0 to 5 months. The data further depict that in each survey year, fever’s prevalence was persistently higher in children living in households where their source of drinking water was unimproved and those who used unclean fuel. Urban residents also had a lower prevalence of fever in under-five children compared to the rural residents and this pattern was observed across the survey rounds.

**Table 2.** Prevalence of Fever by Selected Demographic Characteristics of the Children, per Survey Year.

|  | **2008**<br>(N=22164)<br>n (%) | **2013**<br>(N=26116)<br>n (%) | **2018**<br>(N=11374)<br>n (%) |
| --- | --- | --- | --- |
| **Total** | **3590 (16.2)** | **3483 (13.3)** | **2925 (25.8)** |
| **Characteristics** |  |  |  |
| **Sex of Child** | **4.123\*** | **2.792** | **0.111** |
| Male | 1880 (16.7) | 1803 (13.7) | 1473 (25.6) |
| Female | 1710 (15.7) | 1680 (13.0) | 1452 (25.9) |
| **Child Age (In Months)** | **236.131\*** | **338.382\*** | **149.025\*** |
| 0-5 | 247 (9.3) | 212 (7.5) | 177 (14.7) |
| 6-11 | 514 (19.9) | 541 (17.8) | 372 (30.2) |
| 12-23 | 938 (20.9) | 996 (18.4) | 757 (31.8) |
| 24-35 | 743 (18.0) | 702 (14.3) | 582 (26.6) |
| 36-47 | 625 (14.3) | 575 (11.3) | 547 (25.0) |
| 48-59 | 523 (13.3) | 457 (9.5) | 490 (22.4) |
| **Maternal Age** | **10.013** | **8.509** | **9.537** |
| 15-19 | 221 (18.7) | 180 (14.5) | 127 (28.2) |
| 20-24 | 701 (16.5) | 699 (14.0) | 561 (27.8) |
| 25-29 | 981 (15.4) | 985 (13.5) | 809 (24.9) |
| 30-34 | 759 (15.9) | 739 (12.8) | 652 (24.5) |
| 35-39 | 547 (16.6) | 513 (12.5) | 504 (26.0) |
| 40-44 | 262 (16.5) | 261 (13.4) | 201 (26.0) |
| 45-49 | 119 (17.1) | 106 (14.6) | 71 (24.1) |
| **Maternal Education** | **3.41** | **5.552** | **156.361\*** |
| No Education | 1747 (16.3) | 1659 (13.4) | 1379 (31.4) |
| Primary | 859 (16.5) | 749 (13.9) | 503 (26.6) |
| Secondary | 822 (16.1) | 899 (13.2) | 864 (21.3) |
| Higher | 162 (14.3) | 176 (11.6) | 179 (17.1) |
| **Source of Drinking Water** | **19.767\*** | **4.590\*** | **10.145\*** |
| Improved | 1717 (15.1) | 2004 (13.0) | 1717 (24.7) |
| Unimproved | 1873 (17.3) | 1479 (13.9) | 1208 (27.4) |
| **Cooking Fuel Type** | **7.640\*** | **65.805\*** | **151.739\*** |
| Clean | 461 (14.5) | 399 (9.5) | 358 (15.7) |
| Unclean | 3129 (16.5) | 3084 (14.1) | 2567 (28.2) |
| **Toilet Facility** | **6.048\*** | **0.778** | **55.942\*** |
| Improved | 1651 (15.6) | 1677 (13.1) | 1378 (22.8) |
| Unimproved | 1939 (16.8) | 1806 (13.5) | 1547 (29.0) |
| **Housing Quality Material** | **16.895\*** | **56.614\*** | **197.277\*** |
| Poor | 1737 (17.3) | 1671 (15.2) | 946 (33.7) |
| Average | 477 (15.5) | 512 (12.3) | 596 (30.6) |
| Good | 1376 (15.2) | 1300 (11.9) | 1383 (20.9) |
| **Place of Residence** | **42.322\*** | **4.232\*** | **86.975\*** |
| Urban | 847 (13.6) | 1080 (12.7) | 929 (20.9) |
| Rural | 2743 (17.2) | 2403 (13.6) | 1996 (28.8) |
| **Wealth Index** | **31.415\*** | **64.975\*** | **261.618\*** |
| Poorest | 957 (17.4) | 861 (14.5) | 811 (35.8) |
| Poorer | 886 (16.9) | 883 (14.5) | 668 (29.4) |
| Middle | 721 (16.6) | 773 (14.4) | 634 (25.2) |
| Richer | 603 (15.5) | 580 (11.9) | 503 (21.2) |
| Richest | 423 (13.2) | 386 (10.0) | 309 (15.8) |
| **Region** | **487.378\*** | **740.839\*** | **380.292\*** |
| North Central | 396 (9.4) | 300 (7.6) | 412 (20.5) |
| North East | 1079 (20.5) | 1205 (22.0) | 737 (36.1) |
| North West | 847 (15.4) | 848 (10.2) | 885 (31.7) |
| South East | 514 (25.8) | 479 (20.4) | 371 (23.1) |
| South South | 498 (20.7) | 442 (14.3) | 329 (26.2) |
| South West | 256 (9.2) | 209 (7.1) | 191 (11.4) |

**\*Chi-square value significant at 95% C.I.**

#### Distribution of Short Rapid Breaths

The prevalence of short rapid breath (SRB) among under-five children across the three survey rounds was presented in Table 3. The prevalence of SRB was highest in 2008 (41.7%) and least in 2018 (6.5%). There was a consistently high prevalence of SRB among under-five children living in households where unimproved sources of drinking water were used – the prevalence of SRB were 47.3%, 48.9%, and 7.4% in 2003, 2013, and 2018 respectively compared to households with improved sources of drinking water. SRB prevalence in under-five children was consistently higher in rural areas than in urban. In 2008 and 2013 data for instance, compared to under-five children living in the urban areas where the prevalence of SRB was 36.1% and 32.9%, about 44.0% and 47.6% were found in the rural areas respectively.

**Table 3.** Prevalence of Short Rapid Breaths by Selected Demographic Characteristics, per Year.

|  | 2008<br>(N=2620)<br>n (%) | 2013<br>(N=2605)<br>n (%) | 2018<br>(N=11376)<br>n (%) |
| --- | --- | --- | --- |
| **Total** | **1092 (41.7)** | **1106 (42.5)** | **741 (6.5)** |
| **<u>Characteristics</u>** |  |  |  |
| **Sex of Child** | **0.042** | **0.06** | **0.858** |
| Male | 544 (41.9) | 572 (42.7) | 363 (6.3) |
| Female | 548 (41.5) | 534 (42.2) | 378 (6.7) |
| **Child Age (In Months)** | **7.212** | **12.832\*** | **55.812\*** |
| 0-5 | 107 (42.5) | 86 (42.2) | 67 (5.6) |
| 6-11 | 165 (42.3) | 190 (44.7) | 107 (8.7) |
| 12-23 | 299 (44.6) | 334 (44.3) | 206 (8.6) |
| 24-35 | 202 (37.2) | 233 (45.6) | 138 (6.3) |
| 36-47 | 177 (42.4) | 147 (36.4) | 140 (6.4) |
| 48-59 | 142 (40.8) | 116 (37.8) | 83 (3.8) |
| **Maternal Age** | **20.680\*** | **19.719\*** | **7.356** |
| 15-19 | 72 (47.4) | 63 (45.3) | 28 (6.2) |
| 20-24 | 254 (49.1) | 267 (47.7) | 156 (7.7) |
| 25-29 | 315 (40.8) | 290 (41.0) | 212 (6.5) |
| 30-34 | 216 (37.4) | 230 (39.2) | 169 (6.4) |
| 35-39 | 138 (38.0) | 141 (37.2) | 113 (5.8) |
| 40-44 | 69 (40.1) | 78 (47.3) | 47 (6.1) |
| 45-49 | 28 (42.4) | 37 (54.4) | 16 (5.4) |
| **Maternal Education** | **129.945\*** | **91.417\*** | **22.122\*** |
| No Education | 558 (54.0) | 495 (53.4) | 311 (7.1) |
| Primary | 272 (39.4) | 265 (43.3) | 153 (8.1) |
| Secondary | 235 (31.2) | 295 (33.5) | 231 (5.7) |
| Higher | 27 (19.0) | 51 (27.6) | 46 (4.4) |
| **Source of Drinking Water** | **32.737\*** | **29.953\*** | **10.361\*** |
| Improved | 488 (36.3) | 589 (38.1) | 412 (5.9) |
| Unimproved | 604 (47.3) | 517 (48.9) | 329 (7.4) |
| **Cooking Fuel Type** | **77.870\*** | **113.769\*** | **28.153\*** |
| Clean | 107 (23.3) | 89 (19.8) | 93 (4.1) |
| Unclean | 985 (45.6) | 1017 (47.2) | 648 (7.1) |
| **Toilet Facility** | **17.827\*** | **15.629\*** | **0.723** |
| Improved | 474 (37.5) | 483 (38.5) | 382 (6.3) |
| Unimproved | 618 (45.6) | 623 (46.1) | 359 (6.7) |
| **Housing Quality Material** | **123.180\*** | **111.767\*** | **56.336\*** |
| Poor | 560 (52.6) | 557 (51.8) | 255 (9.1) |
| Average | 173 (47.5) | 196 (50.8) | 149 (7.6) |
| Good | 359 (30.1) | 353 (30.9) | 337 (5.1) |
| **Place of Residence** | **13.815\*** | **52.518\*** | **13.447\*** |
| Urban | 277 (36.1) | 300 (32.9) | 242 (5.5) |
| Rural | 815 (44.0) | 806 (47.6) | 499 (7.2) |
| **Wealth Index** | **141.450\*** | **143.625\*** | **57.612\*** |
| Poorest | 305 (52.2) | 263 (53.0) | 218 (9.6) |
| Poorer | 296 (54.0) | 321 (53.1) | 150 (6.6) |
| Middle | 226 (42.5) | 276 (46.1) | 163 (6.5) |
| Richer | 160 (30.5) | 144 (29.6) | 127 (5.4) |
| Richest | 105 (24.4) | 102 (24.3) | 83 (4.2) |
| **Region** | **228.194\*** | **270.902\*** | **421.444\*** |
| North Central | 143 (41.4) | 149 (46.6) | 78 (3.9) |
| North East | 469 (58.7) | 553 (61.2) | 323 (15.8) |
| North West | 176 (52.9) | 128 (35.9) | 104 (3.7) |
| South East | 112 (27.7) | 132 (36.5) | 102 (6.4) |
| South South | 141 (30.0) | 117 (26.2) | 106 (8.4) |
| South West | 51 (19.0) | 27 (12.5) | 28 (1.7) |

**\*Chi-square values significant at 95% C.I.**

### Optimized Hotspot Analysis of Prevalence of Pneumonia Symptoms

Figs 1-3 show the spatial and temporal distribution of states in Nigeria with a significantly high prevalence of pneumonia symptoms. For Cough, Fig 1 indicates a decrease in states with a significantly high prevalence of cough over time. In 2008, only Gombe has a significantly high prevalence at 90% CI while the other years has no states with significantly high prevalence. Fig 2 indicates an overall decrease in prevalence and states with a significantly high prevalence of fever over time. In 2008, Bayelsa, Rivers, Akwa Ibom, Edo, Anambra, Enugu, Ebonyi and Imo states had significantly high prevalence at 90% CI while in 2013, the south-south states had less significant prevalence which include Delta, Bayelsa and Rivers only at 90% CI. There was no state with a significantly high prevalence of fever in 2018. Fig 3 shows a decrease in the number of states with significantly high prevalence from 2008 to 2018. In 2008, Kaduna, Kano and Jigawa were the states with a significantly high prevalence of short rapid breath. In 2013 and 2018, only Yobe and Taraba has a significantly high prevalence of pneumonia symptoms respectively.

**Fig 1.**
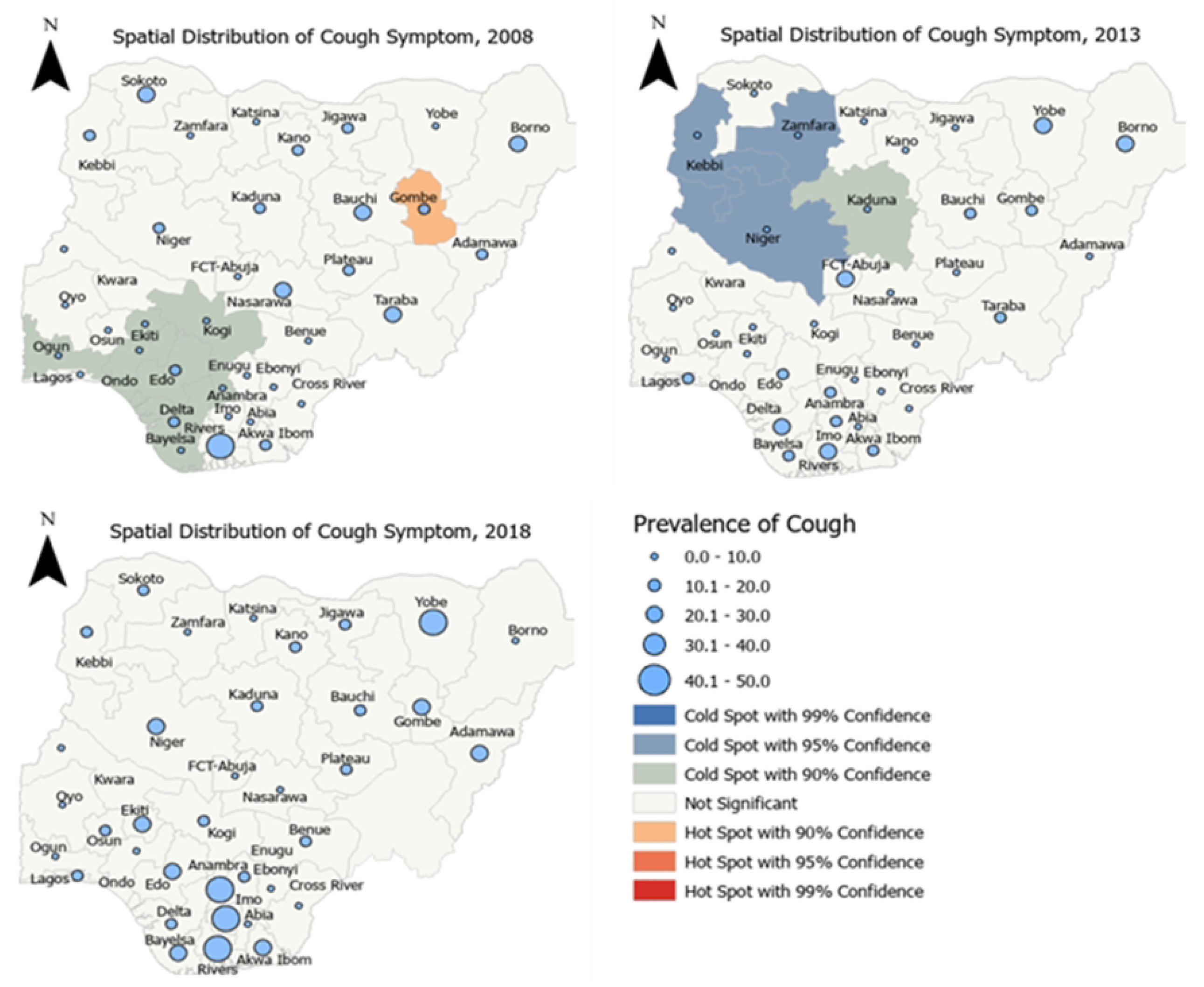
Significance of High Prevalence of Cough in under-five children in Nigeria, 2008-2018.

**Fig 2.**
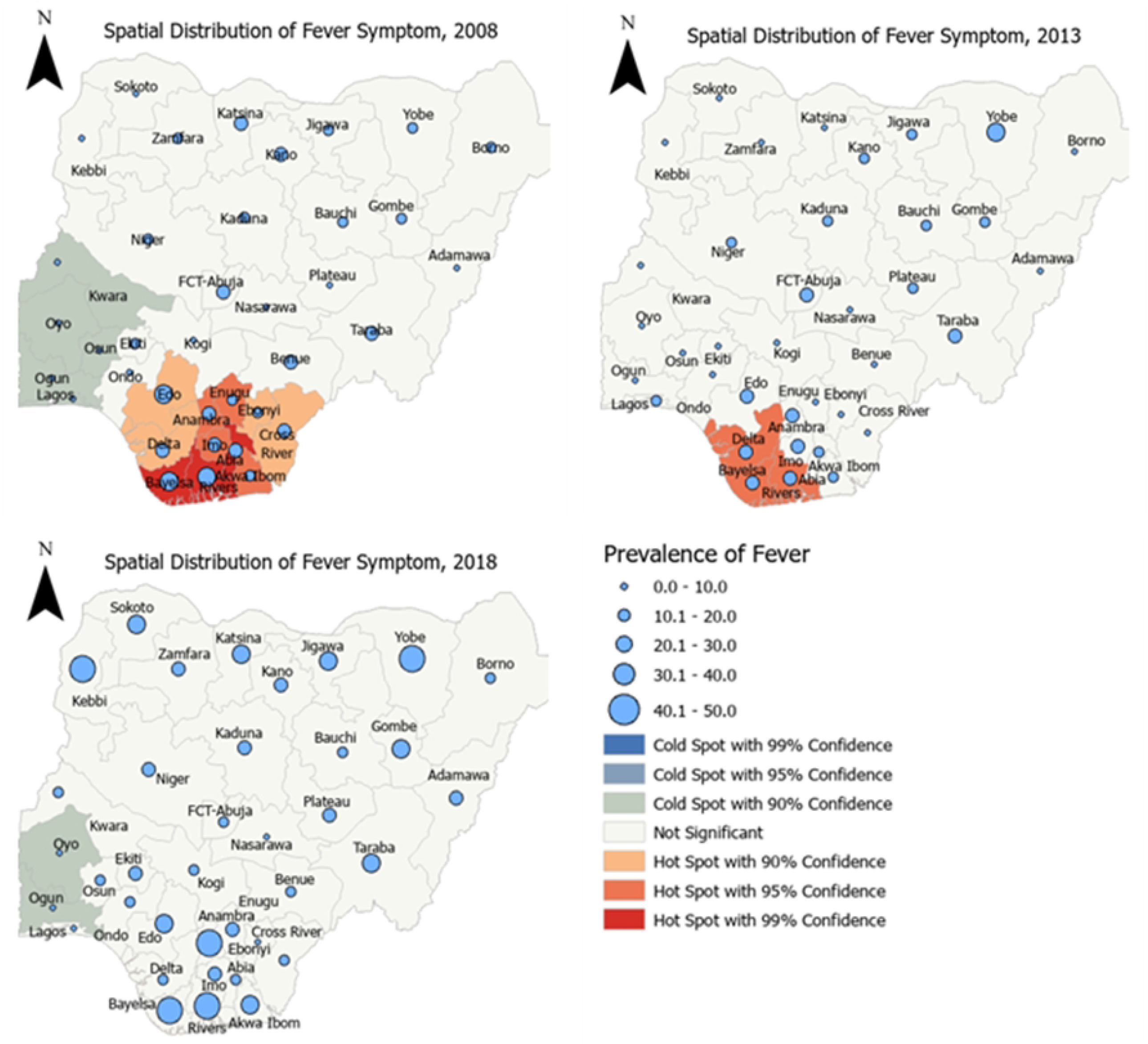
Significance of High Prevalence of Fever in under-five children in Nigeria, 2008-2018.

**Fig 3.**
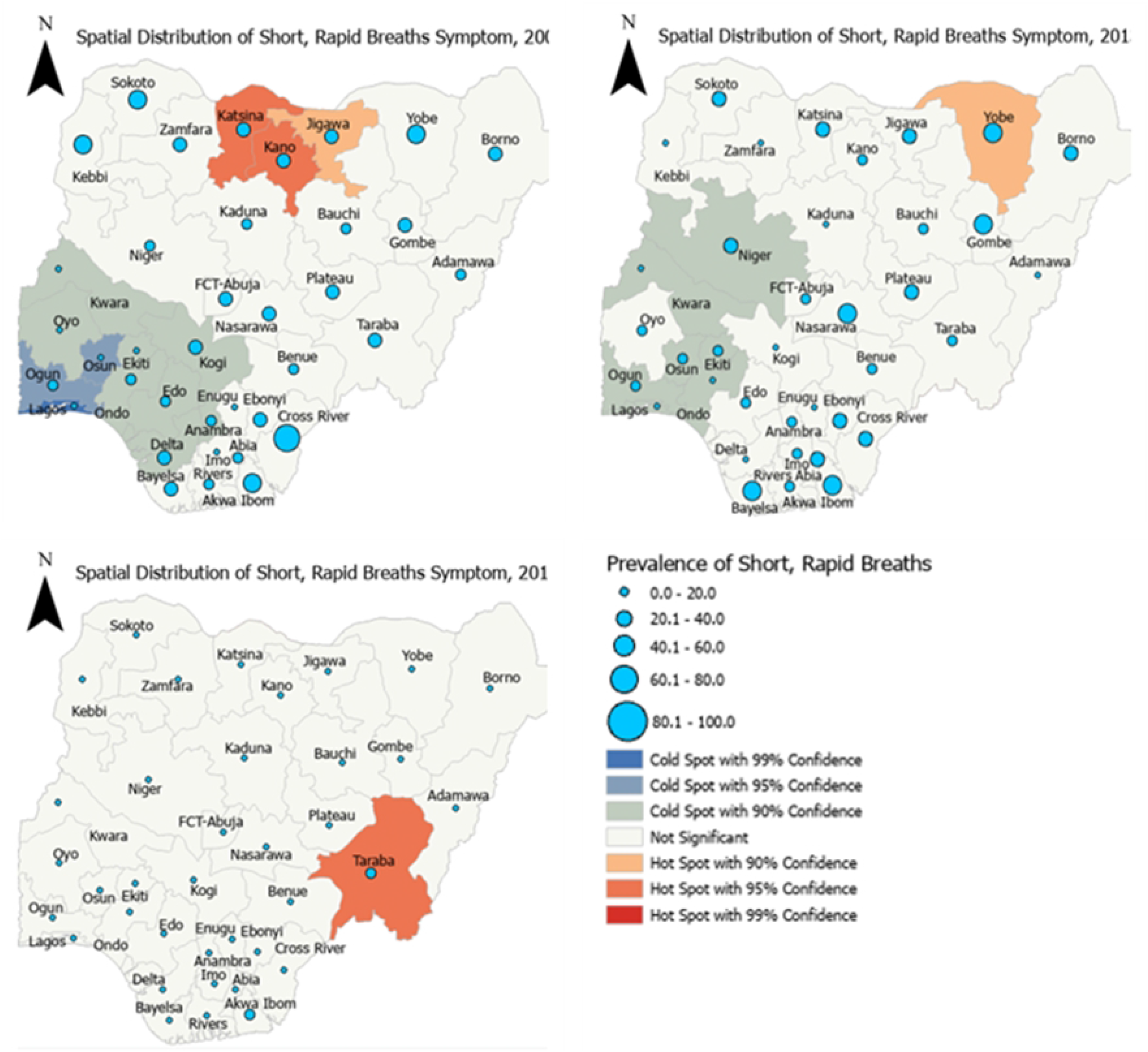
Significance of High Prevalence of Short, Rapid Breaths in under-five children in Nigeria, 2008-2018.

### Factors Associated with Cough, Fever and Short Rapid Breath

Table 4 presents the results of the adjusted MCMC models fitted for each of the symptoms. It is worthy of note that the factors included in these models were put through bivariate analysis to determine their independent association with the respective symptoms before they were seeded into the multiple models. The adjusted odds of cough reduced by 20% (adjusted odds ratio (aOR) =0.80, 95% Credible Interval (CrI): 0.76-0.85) in 2013 but rose by 52% (aOR =1.52, 95% CrI: 1.40, 1.63) in 2018 compared with the likelihood in 2008. Similarly, the adjusted odds of fever reduced by 24% (aOR =0.76, 95% CrI: 0.72, 0.80) in 2013 and nearly doubled (aOR =1.93, 95% CrI: 1.82, 2.05) in 2018. Whereas the adjusted odds of short rapid breaths were insignificant in 2013, but reduced by 91% (aOR =0.09, 95% CrI: 0.08, 0.11) in 2018 compared with 2008.

**Table 4.**
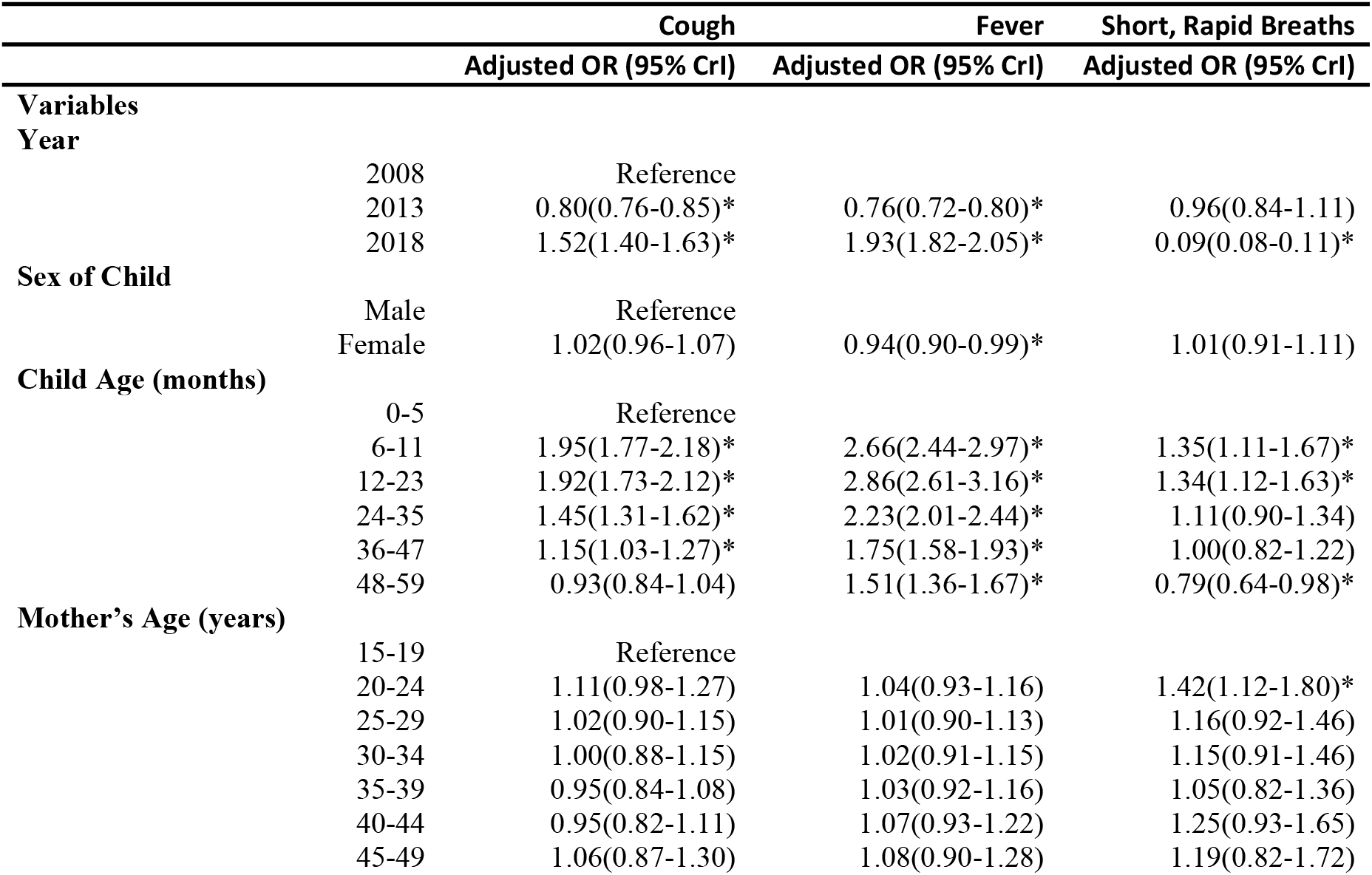
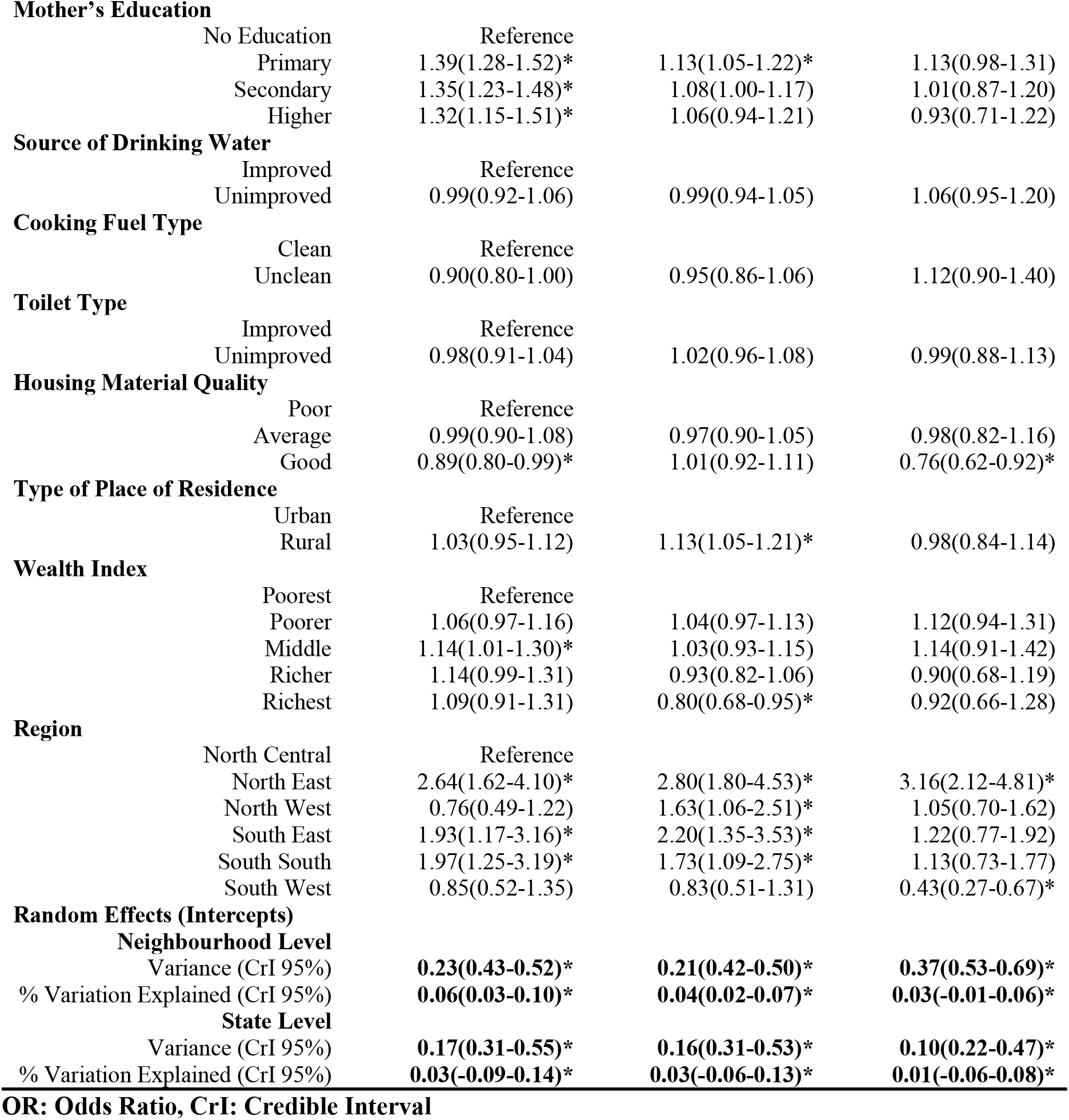
Results of the Monte Carlo Markov Chain Models for each Symptom of Pneumonia.

|  |  | Cough | Fever | Short, Rapid Breaths |
| --- | --- | --- | --- | --- |
|  |  | Adjusted OR (95% CrI) | Adjusted OR (95% CrI) | Adjusted OR (95% CrI) |
| **Variables** |  |  |  |  |
| **Year** | 2008 | Reference |  |  |
|  | 2013 | 0.80(0.76-0.85)* | 0.76(0.72-0.80)* | 0.96(0.84-1.11) |
|  | 2018 | 1.52(1.40-1.63)* | 1.93(1.82-2.05)* | 0.09(0.08-0.11)* |
| **Sex of Child** | Male | Reference |  |  |
|  | Female | 1.02(0.96-1.07) | 0.94(0.90-0.99)* | 1.01(0.91-1.11) |
| **Child Age (months)** | 0-5 | Reference |  |  |
|  | 6-11 | 1.95(1.77-2.18)* | 2.66(2.44-2.97)* | 1.35(1.11-1.67)* |
|  | 12-23 | 1.92(1.73-2.12)* | 2.86(2.61-3.16)* | 1.34(1.12-1.63)* |
|  | 24-35 | 1.45(1.31-1.62)* | 2.23(2.01-2.44)* | 1.11(0.90-1.34) |
|  | 36-47 | 1.15(1.03-1.27)* | 1.75(1.58-1.93)* | 1.00(0.82-1.22) |
|  | 48-59 | 0.93(0.84-1.04) | 1.51(1.36-1.67)* | 0.79(0.64-0.98)* |
| **Mother's Age (years)** | 15-19 | Reference |  |  |
|  | 20-24 | 1.11(0.98-1.27) | 1.04(0.93-1.16) | 1.42(1.12-1.80)* |
|  | 25-29 | 1.02(0.90-1.15) | 1.01(0.90-1.13) | 1.16(0.92-1.46) |
|  | 30-34 | 1.00(0.88-1.15) | 1.02(0.91-1.15) | 1.15(0.91-1.46) |
|  | 35-39 | 0.95(0.84-1.08) | 1.03(0.92-1.16) | 1.05(0.82-1.36) |
|  | 40-44 | 0.95(0.82-1.11) | 1.07(0.93-1.22) | 1.25(0.93-1.65) |
|  | 45-49 | 1.06(0.87-1.30) | 1.08(0.90-1.28) | 1.19(0.82-1.72) |

**Mother's Education**
|  |  |  |  |
| --- | --- | --- | --- |
| No Education | Reference |  |  |
| Primary | 1.39(1.28-1.52)* | 1.13(1.05-1.22)* | 1.13(0.98-1.31) |
| Secondary | 1.35(1.23-1.48)* | 1.08(1.00-1.17) | 1.01(0.87-1.20) |
| Higher | 1.32(1.15-1.51)* | 1.06(0.94-1.21) | 0.93(0.71-1.22) |

**Source of Drinking Water**
|  |  |  |  |
| --- | --- | --- | --- |
| Improved | Reference |  |  |
| Unimproved | 0.99(0.92-1.06) | 0.99(0.94-1.05) | 1.06(0.95-1.20) |

**Cooking Fuel Type**
|  |  |  |  |
| --- | --- | --- | --- |
| Clean | Reference |  |  |
| Unclean | 0.90(0.80-1.00) | 0.95(0.86-1.06) | 1.12(0.90-1.40) |

**Toilet Type**
|  |  |  |  |
| --- | --- | --- | --- |
| Improved | Reference |  |  |
| Unimproved | 0.98(0.91-1.04) | 1.02(0.96-1.08) | 0.99(0.88-1.13) |

**Housing Material Quality**
|  |  |  |  |
| --- | --- | --- | --- |
| Poor | Reference |  |  |
| Average | 0.99(0.90-1.08) | 0.97(0.90-1.05) | 0.98(0.82-1.16) |
| Good | 0.89(0.80-0.99)* | 1.01(0.92-1.11) | 0.76(0.62-0.92)* |

**Type of Place of Residence**
|  |  |  |  |
| --- | --- | --- | --- |
| Urban | Reference |  |  |
| Rural | 1.03(0.95-1.12) | 1.13(1.05-1.21)* | 0.98(0.84-1.14) |

**Wealth Index**
|  |  |  |  |
| --- | --- | --- | --- |
| Poorest | Reference |  |  |
| Poorer | 1.06(0.97-1.16) | 1.04(0.97-1.13) | 1.12(0.94-1.31) |
| Middle | 1.14(1.01-1.30)* | 1.03(0.93-1.15) | 1.14(0.91-1.42) |
| Richer | 1.14(0.99-1.31) | 0.93(0.82-1.06) | 0.90(0.68-1.19) |
| Richest | 1.09(0.91-1.31) | 0.80(0.68-0.95)* | 0.92(0.66-1.28) |

**Region**
|  |  |  |  |
| --- | --- | --- | --- |
| North Central | Reference |  |  |
| North East | 2.64(1.62-4.10)* | 2.80(1.80-4.53)* | 3.16(2.12-4.81)* |
| North West | 0.76(0.49-1.22) | 1.63(1.06-2.51)* | 1.05(0.70-1.62) |
| South East | 1.93(1.17-3.16)* | 2.20(1.35-3.53)* | 1.22(0.77-1.92) |
| South South | 1.97(1.25-3.19)* | 1.73(1.09-2.75)* | 1.13(0.73-1.77) |
| South West | 0.85(0.52-1.35) | 0.83(0.51-1.31) | 0.43(0.27-0.67)* |

**Random Effects (Intercepts)****Neighbourhood Level**
|  |  |  |  |
| --- | --- | --- | --- |
| Variance (CrI 95%) | **0.23(0.43-0.52)\*** | **0.21(0.42-0.50)\*** | **0.37(0.53-0.69)\*** |
| % Variation Explained (CrI 95%) | **0.06(0.03-0.10)\*** | **0.04(0.02-0.07)\*** | **0.03(-0.01-0.06)\*** |

**State Level**
|  |  |  |  |
| --- | --- | --- | --- |
| Variance (CrI 95%) | **0.17(0.31-0.55)\*** | **0.16(0.31-0.53)\*** | **0.10(0.22-0.47)\*** |
| % Variation Explained (CrI 95%) | **0.03(-0.09-0.14)\*** | **0.03(-0.06-0.13)\*** | **0.01(-0.06-0.08)\*** |

**OR: Odds Ratio, CrI: Credible Interval**

## Discussion

This study was designed to identify and explain the Spatio-temporal distribution and risk factors associated with pneumonia symptoms among under-five children in Nigeria using the Markov Chain Monte Carlo Random Intercept Logistic Regression. The study was conceptualized against the backdrop and gap observed in similar studies previously conducted in Nigeria. A temporal pattern was observed in the model. This corroborated the need for spatial analyses of the symptoms by survey years. The likelihood of cough and fever both reduced between 2008 and 2013 but the odds rose by 52% and 93% respectively in 2018. In sharp contrast, the adjusted odds of short rapid breaths were reduced by 91% in 2018 compared with 2013. The temporal pattern observed in the current study corroborates findings from similar studies conducted elsewhere [43,44].

The in-depth spatial hotspot analysis indicated a significant reduction in the prevalence of the symptoms of pneumonia from 2008 to 2018. This decrease may not be unconnected with the various interventional programme and efforts of the government of Nigeria in conjunction with the United Nations and other international partners. For instance, the introduction of the National Pneumonia Control Strategy and Implementation Plan (NPCSIP) by the Nigerian government in the year 2019 towards reducing the high number of childhood mortality due to pneumonia and the menace of pneumonia-related morbidities/deaths in under-five children [45] can explain the reason for our finding. The impact of the government’ interventions may be responsible for the statistically significant reduction of these symptoms over the years. In our study, the gender of the child was found to be related to fever. Gender has been said to be a significant epidemiological factor for most diseases [46]. It is important to note that the males also had a higher prevalence of fever as a symptom of pneumonia for all the years under study. Similarly, Victoria et al. found gender to be significantly related to pneumonia but was expressed as the males having a higher odds of experiencing pneumonia, though the odds of a male having pneumonia than a female in this study was lower [47]. The role of sex hormones in the immunity of individuals may have been responsible for this difference [46].

Children aged 6 to 47 months had higher odds of experiencing cough, fever, or short rapid breaths as symptoms of pneumonia relative to 0 to 5 months children. Previous studies also reported that the odds of contracting pneumonia was higher among children aged 6 to 48 months and lower among children aged 48 to 59 months [31]. This confirms the assertion that younger age corresponds to higher odds of contracting pneumonia in children [7]. Maternal age was significant only for short rapid breaths in mothers aged 20 to 24 where they had higher odds of bringing about short rapid breaths as a pneumonia symptom in their children. This may be due to inexperience in child care or their lack of knowledge or perception of pneumonia. Another study that examined how age affects mothers’ perception and knowledge of pneumonia in children found that women aged 18-24 were grossly lacking in knowledge [48]. This poor knowledge may have accounted for mothers aged 20-24 ability to prevent the occurrence of pneumonia (short rapid breaths) in under-five children. Maternal education goes to a large extent in affecting their knowledge of pneumonia and its preventive measures. It is expected higher education among mothers would correspond to lower odds of the occurrence of pneumonia symptoms in their children. However, this may not necessarily be the case, as there may be a difference between knowledge and practice, particularly regarding health outcomes [49].

The sources of drinking water were not found to be significant for the three symptoms of pneumonia in this study contrary to earlier findings [50,51] despite a large-scale lack of quality water in Nigeria [52,53]. Unclean cooking fuel was found not to be significantly associated with cough, fever, and short rapid breaths as symptoms of pneumonia. This is against the expectation of the effect of pollution caused by unclean solid fuels. The findings were corroborated in a study of the household environment and symptoms of childhood acute respiratory tract infections in Nigeria [54].

The role that housing plays in the state of the health of the people cannot be overlooked and overemphasized [55]. Homes that are healthy with enough quality ventilation and insulation guarantees the health of the members of such households, free from pests and contaminant [56]. Findings from this study showed that the odds of cough and short rapid breaths as symptoms of pneumonia were lower among children residing in buildings with good housing material quality. Overcrowding, poor sleeping arrangements, and poor ventilation dominate most housing with poor quality in Nigeria, but findings on toilet facility contrasts previous literature that sanitation and hygiene is a major contribution to childhood pneumonia [57]. A possible basis for the disparity in the odds of the symptoms across the geopolitical regions can be attributed to variations in social and economic development in these regions [54,58,59].

## Study Limitations and Strengths

The use of secondary data for this study limits the choice of variables we explored. The data might have suffered recall bias as this study was a cross-sectional study wherein respondents were made to recall past events without any means of verification by the data collectors. Notwithstanding, the data collection procedures and methods adopted by DHS minimized the effect of such bias. The DHS is well-known for its high quality as a result of the data collection approach used by the data originators. The data used is also nationally representative data covering nearly two decades. Also, the pneumonia symptoms available in the DHS dataset are not exhaustive and prevented the measurement of pneumonia from a diagnostic perspective which would have enabled us to have a variable measuring pneumonia as a disease. A major strength of our study is the assessment of factors associated with individual symptoms compared to other studies that studied their composite. The use of Bayesian models in place of frequentist models in understanding the associated risk factors helped to avoid the problems of the inconsistency of results associated with the frequentist approach.

## Conclusion

This study provided evidence that variations exisits in the prevalence of pneumonia symptoms among under-five children across the states in Nigeria. The northern states were more characterized by *short, rapid breath*s while the southern states were more characterized by *fever* but both regions have a fair share of *cough* among under-five children.

For the cough and fever symptoms, the increased prevalence between 2013 and 2018 indicates that the symptoms need more attention. Although care should be taken in interpreting the significance of a factor to at least one of the symptoms of pneumonia as significance to pneumonia, our findings supported positive change in pneumonia prevention by providing evidence of risk factors influencing the symptoms of pneumonia. Child’s age and region of residence were significant determinants of all pneumonia symptoms considered in the study. While place of residence was a significant predictor of fever, wealth and mother’s education was significant for cough and fever.

## Recommendations

Government and other relevant stakeholders should do more in the area of providing good housing conditions to its population as this will alleviate the prevalence of pneumonia symptoms in Nigeria. The interventions for pneumonia control towards the United Nation’s Millennium Development Goal (MDG) 4 of reducing child mortality by two-thirds should be sustained and improved upon since these strategies have yielded good results. Effective case management and treatment at the community level should be ensured. The high prevalence of pneumonia symptoms in Nigeria as evidenced in this study points to the need for government to improve on the existing sensitization frameworks on home prevention of pneumonia symptoms in under-five children in Nigeria.

## Data Availability

The data underlying the results presented in the study are available from www.dhsprogram.com/data

## Declarations

### Ethics approval and consent to participate

The owners of the secondary data used in this analysis obtained the necessary ethical approval and consent prior to data collection. There was no need for further approvals on our part as open-source secondary data.

### Authors’ contributions

AKA, LTV and FAF conceptualized and designed the study. AKA and LTV analyzed and interpreted the data while FAF and AAS were senior authors overseeing the analysis and interpretation of the results. AKA and LTV drafted the manuscript FAF and AAS reviewed and edited the manuscript. All authors have read and agreed to the published version of the manuscript.

## Acknowledgements

The authors are grateful to ICF Macro, USA, for granting the authors the request to use the Demographic and Health Survey data.

## Supporting Information

**S1 Table. Distribution of demographic characteristics for cough per year of survey**

**S2 Table. Distribution of demographic characteristics for fever per year of survey**

**S3 Table. Distribution of demographic characteristics for short rapid breaths per year of survey**

**S4 Table. Prevalence of cough by state and year of survey**

**S5 Table. Prevalence of fever by state and year of survey**

**S6 Table. Prevalence of short rapid breaths symptoms by state and year of survey**

